# Differentiating kinds of systemic chronic stressors with relation to psychotic-like experiences in late childhood and early adolescence: the stimulation, discrepancy, and deprivation model of psychosis

**DOI:** 10.1101/2020.08.05.20168492

**Authors:** Teresa Vargas, Katherine S.F. Damme, K. Juston Osborne, Vijay A. Mittal

## Abstract

**INTRODUCTION:** Chronic stress exposure occurs at the systems level, and is a key etiological factor in the development of psychotic disorders. However, conceptualizations distinguishing the impact of distinct dimensions of stress exposure are lacking; further, the magnitude of effect for differing exposures has yet to be explored.

**METHODS:** Exploratory factor analysis was conducted to distinguish domains of environmental exposures in a nationally representative sample of 7,446 youth from the Adolescent Brain Cognitive Development (ABCD) study. Environmental exposures were associated to psychotic-like experiences (PLEs). The magnitude of associations was compared among different environmental exposures. As an exploratory aim, objective versus subjective measures of environmental risk exposure were compared.

**RESULTS:** Six factors were defined, four of which fit theoretically with the Stimulation Discrepancy, and Deprivation (SDD) theory of developmental stress exposure and psychosis. The three domains of *stimulation* (high attentional demands and lack of safety), *discrepancy* (low social capital, social exclusion and lack of belonging), and *deprivation* (lack of developmentally appropriate environmental enrichment) were associated with PLEs, as predicted. Compared to exposures in other domains, exposures in the *deprivation* domain exhibited a significantly stronger association with PLEs. Objective and subjective measures converged in direction of association, though self-report *stimulation* exhibited a significantly stronger association with PLEs compared to objective *stimulation* domain measures.

**DISCUSSION:** Current results suggest considering distinct environmental exposures as they relate to psychosis liability could inform putative mechanisms and degrees of vulnerability. The approach offers a valuable perspective to health policy efforts aimed at psychopathology prevention and intervention efforts.

## Introduction

Chronic stress has been widely implicated to play a causal role in the etiology of psychosis^1-5^. A wide array of work has examined individual-level chronic stressors and their relation to psychosis risk. More recently, however, the field has looked toward examining systems-level factors, such as neighborhood features. A recent review synthesizing the literature on systemic environmental risk factors for psychotic disorders hypothesized that exposures fall within three domains. These hypothesized environmental risk domains include *stimulation* (lack of safety and high attentional demands), *discrepancy* (lack of belonging and social exclusion) and *deprivation* (lack of needed environmental enrichment). To date, relevant systemic environmental risk literature has primarily focused on adult populations, whereas late childhood and early adolescent periods could represent a highly informative timespan for early detection and prevention efforts^6-10^. Further, while a body of literature supports the important roles for each of these three domains, they have yet to be directly tested together. The present investigation directly tested this theory in two stages. First by examining whether risk factor exposures would separate into the hypothesized domains. Then relating the resulting domains to psychotic-like experiences (PLEs) to assess relevance to psychosis risk. Finally, by comparing the magnitude of effects for observed associations by domain, in order to explore differences in degrees of vulnerability. Examining chronic stress both through an individual and systems level perspective by synthesizing these factors into environmental domains of influence is a critical priority. Despite the considerable research attention dedicated to this class of disorder, the field also lacks a clear understanding of environment x liability interactions, particularly in the pediatric developmental periods. Given the challenging prognosis, a better understanding of causal factors, and individual and systems level prevention efforts are paramount.

Psychotic disorders are chronic in nature, difficult to treat, and highly debilitating, constituting one of the top 15 leading causes of disability worldwide^11^. After psychotic illness onset, confounds related to factors such as medication use, social and functional decline make it difficult to distill factors driving illness onset. As such, assessment of associated symptoms or experiences on the psychosis spectrum, such as PLEs provides a promising alternative for identifying factors relating to psychosis etiology^12^. PLEs, including experiences such as unusual beliefs, suspiciousness, and perceptual abnormalities, have been associated with pathogenic factors implicated in formal psychosis^6-10^. Further, PLEs are experienced by 13-15% of children^13,14^, and childhood experiences of PLEs have been shown to increase later risk for psychotic disorder onset ^14-16^. Investigating PLEs in childhood provides an opportunity to understand environmental risk factors early in development, prior to illness related confounds. PLEs may also be of key importance during a critical developmental period in which prevention could be particularly effective.

Chronic stress exposure is a central contributing factor in the development of psychotic disorders^17,18^. Robust and varied evidence suggests a wide host of stressors can cumulatively contribute to psychosis risk^1-5^. In the neural diathesis stress model, for example, posits that the accumulation of stressors can comprise “multiple hits” which acting on a vulnerable system can increase likelihood of developing a psychotic disorder^4,19^. Although this theory highlights the importance of conceptualizing stressors collectively, isolating qualitatively distinct stressors could also be uniquely informative to psychosis etiology. Work on trauma exposure, for instance, has illuminated differing neurodevelopmental consequences depending on the type of trauma^20-22^. Beyond trauma, distinct domains of chronic stressors could contribute to risk for developing a psychotic disorder. Additionally, these domains could aid in the understanding the complex and multifaceted nature of the illness. Isolating the impacts of differing dimensions of experience could be informative to psychotic disorder etiology. Indeed the non-psychiatric literature has successfully spearheaded conceptualizing neurodevelopment in this manner^22^. Despite this fact, the psychotic disorder literature has largely conceptualized chronic stressors collectively, with less attention given to understanding specific kinds of exposures.

As individuals we operate within a larger environmental, social and cultural context (i.e. structural factors); as such, chronic stress can occur at the systems level^23,24^. Local structural characteristics (such as neighborhood socioeconomic status or cultural integration) could systematically affect well-being and risk for psychopathology. Increasing understanding of structural barriers to mental health has strong potential to inform health policy initiatives, along with prevention and intervention efforts at the societal level. To this end, our group has developed a literature-backed theoretical model of different types of structural exposures, along with distinct intermediary mechanisms of impact and proposed relevant neural systems^25^—the Stimulation Discrepancy Deprivation (SDD) model of psychosis. The *stimulation* domain includes crime exposure at the neighborhood level^26^, along with population density^27^,^28^, with intermediary mechanisms of lack of safety^28^,^29^ and high attentional demands^30^,^31^. The *discrepancy* domain, on the other hand, includes environmental and cultural characteristics conferring a lack of belonging and social exclusion^32^-^39^. Lastly, the *deprivation* domain constitutes environments lacking socioeconomic, educational, or material resources^40-43^, with intermediary mechanisms of lack of necessary environmental enrichment^44,45^. Each domain was theorized to relate to psychosis risk through both common (consistent with the diathesis stress model) and distinct mechanisms. While each domain benefits from support from the psychosis literature, the distinctiveness of the domains has yet to be tested as they relate to the psychosis continuum. The degree to which each domain contributes to psychosis risk is unclear.

Understanding each domain as it relates to degrees of risk for psychopathology could be immensely useful in identifying and prioritizing treatment targets. Conceptualizing degrees of risk could also inform etiological models of psychotic disorders. Further, existing evidence in support of each domain has often honed in on adolescent or young adult populations. Determining whether structural environmental risk exposures are also impactful earlier in development (during childhood and early adolescence), and with regards to sub-threshold psychotic symptoms, would lend granularity to our understanding of environmental risk exposure across developmental periods.

The current investigation utilized a large nationally representative sample of youth aged 9 to 11 years old to further understand exposure to environmental chronic stressors in relation to PLEs. The first aim was to directly test the SDD theory by exploring whether relevant items would load into factors consistent with the 3 hypothesized domains. The second aim was to determine whether the environmental chronic stress domains would relate to PLEs, consistent with the SDD theory. The third aim was to then compare relative strengths of existing associations between environmental exposures and PLEs (in order to see whether certain exposures would show greater associations with psychosis risk than others). A final exploratory aim sought to determine whether associations between objective, Census derived neighborhood metrics of environmental exposures and self-report measures indexing the same exposure would exhibit relations similar in magnitude.

## Methods

### Participants

The ABCD dataset includes a large representative sample of children aged 9-11 years old across 21 centers in the United States ^46,47^ All centers obtained the parents’ informed consent as well as the children’s assent. Research procedures followed ethical guidelines laid out by respective Institutional Review Boards (doi: 10.15154/1519171). The current sample used baseline data, and constituted 7446 participants (who had available data for items in the final factor solution, as well as PLE data). A subset of 7,385 were used for the EFA, since they had available data on all self-report items that were initially considered for factor analysis, and a subset of 6994 also had available data on “objective” neighborhood measures, PLEs, and self-report items used in the factor analysis.

### PLEs

The Prodromal Questionnaire-Brief Child version was used to assess psychotic-like experiences^48-50^. The 21-item self-report questionnaire has been previously validated in the ABCD study sample^48^. The questionnaire asked participants about specific PLEs that were endorsed with a binary response (yes/no). Participants also indicated whether there was distress related to endorsed symptoms on a visual response 5-item Likert scale. Consistent with prior research, distress scores were calculated whereby the total number of endorsed symptoms were weighed by level of distress (0 indicates zero endorsement, 1 indicates endorsement without distress, and 2-6 indicate endorsement with incremental distress levels)^48,49^.

### Self-report questionnaires

Self-report scales relevant to theoretical interest in three domains of deprivation, discrepancy, and stimulation were chosen across numerous administered scales^25^. These included the ABCD Parent Multi-Group Ethnic Identity-Revised Survey (MEIM)^51^, which separates into “ethnic identity search” and “affirmation, belonging and commitment”^51^. The ABCD Parent Vancouver Index of Acculturation (VIA)—Short Survey^52^; which subdivides into “heritage” and “American” subscores^52^. Other scales administered included the ABCD Youth Neighborhood Safety/Crime Survey Modified from PhenX (NSC), ABCD Parent neighborhood safety/crime survey modified from PhenX (NSC)^53,54^, ABCD Parent Acculturation Survey Modified from PhenX (ACC), ABCD Youth Acculturation Survey Modified from PhenX (ACC)^55,56^, and the ABCD Parents Demographics survey,. Primary guardians/parents of the youth completed the ABCD Parent MEIM, ABCD Parent neighborhood safety/crime survey modified from PhenX (NSC), ABCD Parent VIA, and ABCD Parent ACC. Youth completed the ABCD Youth NSC, as well as the ABCD Youth ACC.

### Objective neighborhood features

Residential history was collected through addresses where participants had lived across their lifetime. Addresses were used to determine Census tracts corresponding to each location. Each tract represents Census-delineated neighborhoods. Census and Federal Bureau of Investigation (FBI) data was used to calculate neighborhood population density, total crimes occurring in certain neighborhood, and the area deprivation index (ADI). The ADI metric has been successfully adapted to measure neighborhood deprivation; it is calculated based on the American Community Survey 2015 5-year summary ^57^ Since these metrics are compiled based on government data, they will be referred to as “objective neighborhood features,” drawing a contrast from neighborhood features of interest that are also assessed through self-report (such as the youth and adult NSC).

### Exploratoryfactoranalysis

To determine whether environmental risk factors would fall within hypothesized domains^25^, an exploratory factor analysis was conducted on the self-report scales with the minimum residuals method^58^ using the “psych” package in r^59^. Given the theoretical expectation that some factors would correlate, an oblimin rotation was chosen. Number of factors were decided based on inspection of the scree plot, as well as based on theoretical consistency and interpretability. A cut-off value of 0.4 was chosen for factor loadings; items falling beneath this threshold were excluded^60^.

### Associations between self-report factors, neighborhood features and PLEs

To determine whether PLEs and self-report factors related to each other, Spearman correlations were conducted adjusting for age and sex. Spearman correlations adjusting for age and sex were also conducted to test the association between PLEs and objective neighborhood features. A central aim was to compare the strength of associations between psychotic like experiences and self-report items, as well as between objective neighborhood features and psychotic-like experiences. Differences between correlations between PLEs, self-report and objective neighborhood features indexing the same construct were also tested (i.e. self-report neighborhood safety and total number of crimes in neighborhood, neighborhood deprivation index and self-reported lack of resources). To test whether associations were significantly different, correlation coefficients were converted into a z-score using Fisher’s r-to-z transformation^61^. Then, the asymptotic covariance of the estimates were computed, and used in an asymptotic z-test to determine whether one correlation was significantly greater than the other^62,63^. All analyses were Bonferroni corrected by dividing α = 0.05 by the number of tests conducted^64,65^.

## Results

### Exploratory factor analysis for self-report items

The Kaiser-Meyer-Olkin (KMO) and Bartlett’s test of sphericity indicated the data was appropriate to analyze. The KMO measure of sampling adequacy was 0.89^66-68^. Bartlett’s test of sphericity was significant, χ^2^(465) = 137060.4, p<0.05^69^. The six-factor solution explained 51% of the variance, and was chosen (a) after inspection of the screeplot, as well as (b) based on previous theoretical support, and (c) considering that seven and eight factor solutions were difficult to interpret and had an insufficient number of primary loadings. Five items were eliminated from the original variable set due to failure to meet minimum criteria of having a primary factor loading of 0.4 or above, with 3 of the items also not contributing to the factor structure. The self-report items completed by the youth did not load onto the factor structure (“How well do you speak English?” and “My neighborhood is safe from crime”). One parent reported item did not load onto the factor structure (“Besides English, do you speak or understand another language or dialect?”). Two items (parent reported “How well do you speak English?” and “In the past 12 months, were evicted from your home for not paying the rent or mortgage”) did not meet the requirement of having a factor loading of 0.4 or above.

After removing these five items, the six-factor solution exploratory factor analysis solution using minimum residuals with an oblimin rotation explained 58% of the variance. All items in the analysis had primary loadings over 0.4 (Table 2). With regards to reliability, across the entire dataset, ω _total_ = 0.94 ^70^. Reliability was adequate across factors as well (factor 1, ω _total_ = 0.91; factor 2, ω _total_ = 0.85; factor 3, ω _total_ = 0.90; factor 4, ω _total_ = 0.89; factor 5, ω _total_ = 0.75; factor 6, ω _total_ = 0.75).

**Table 1.**
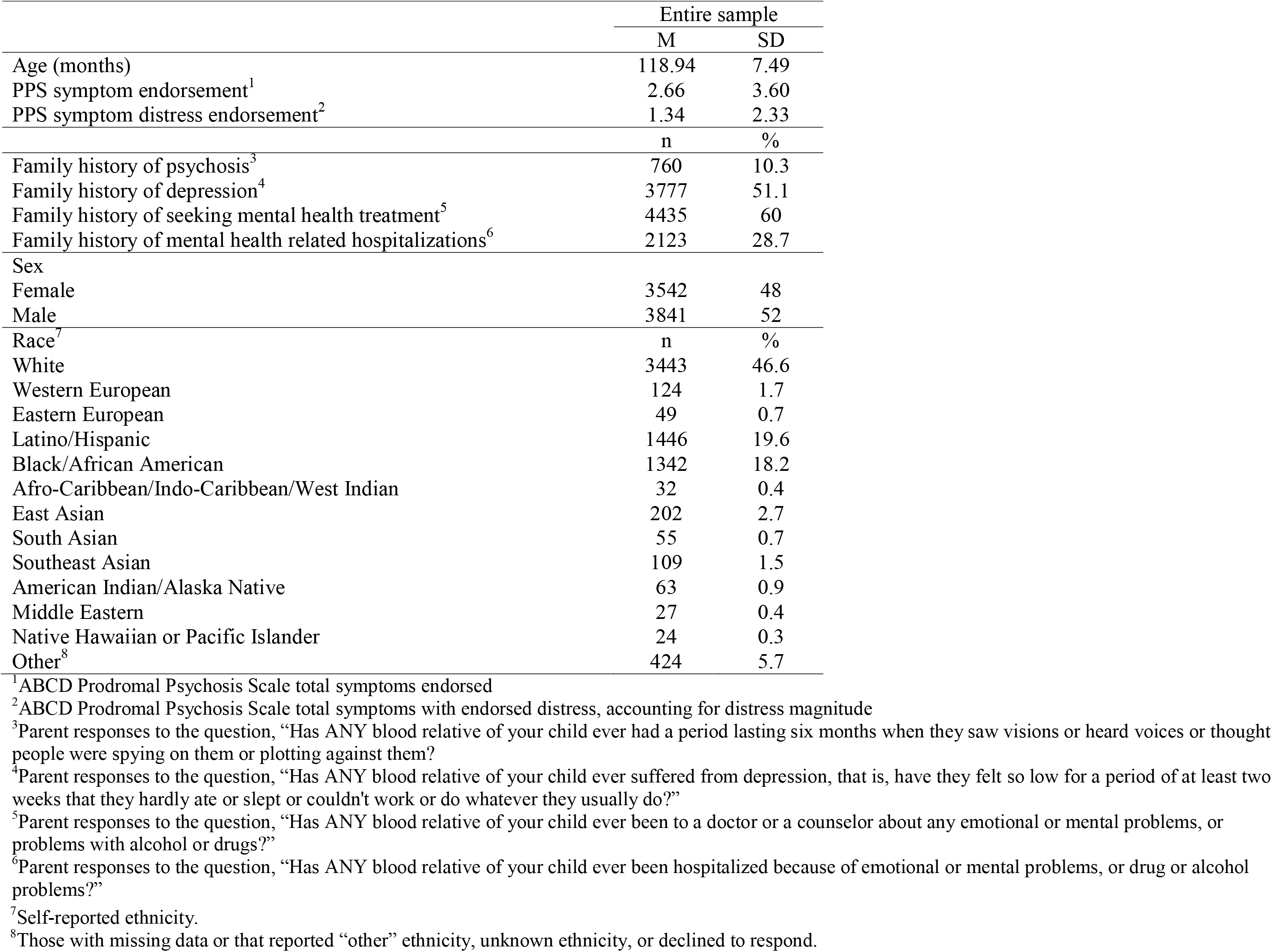
Demographic characteristics.

**Table 2.**
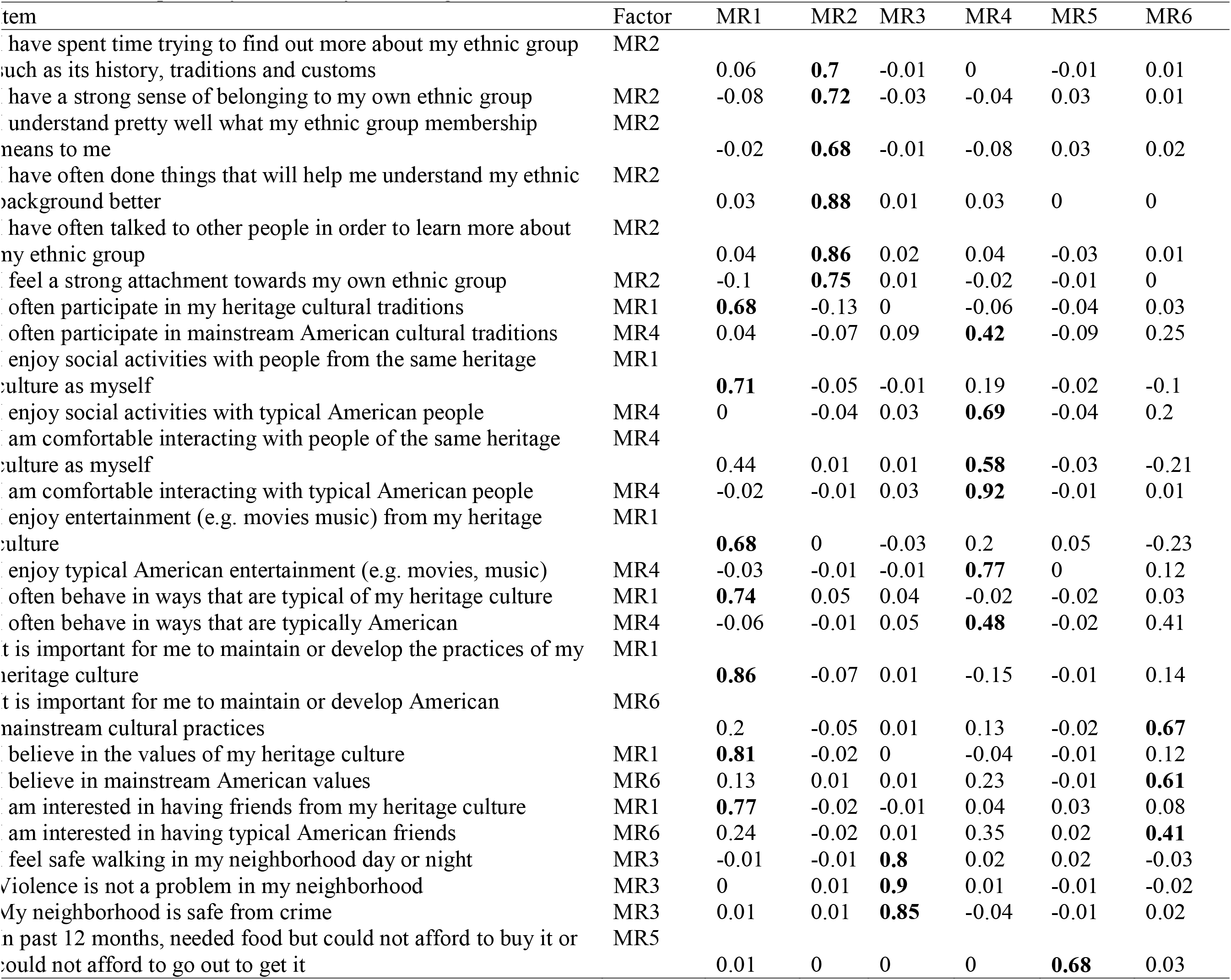
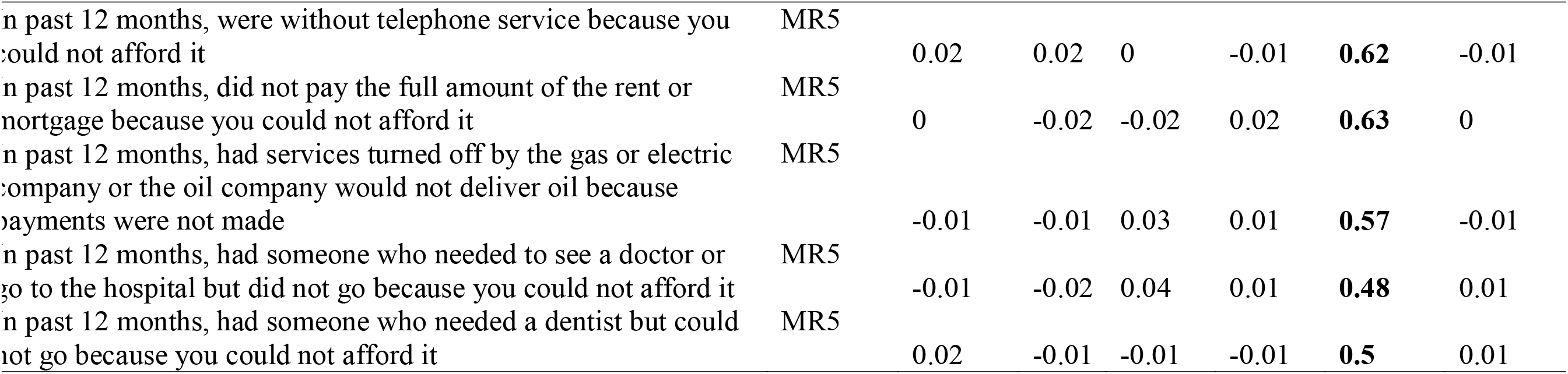
Exploratory factor analysis loadings.

**Table 3.**
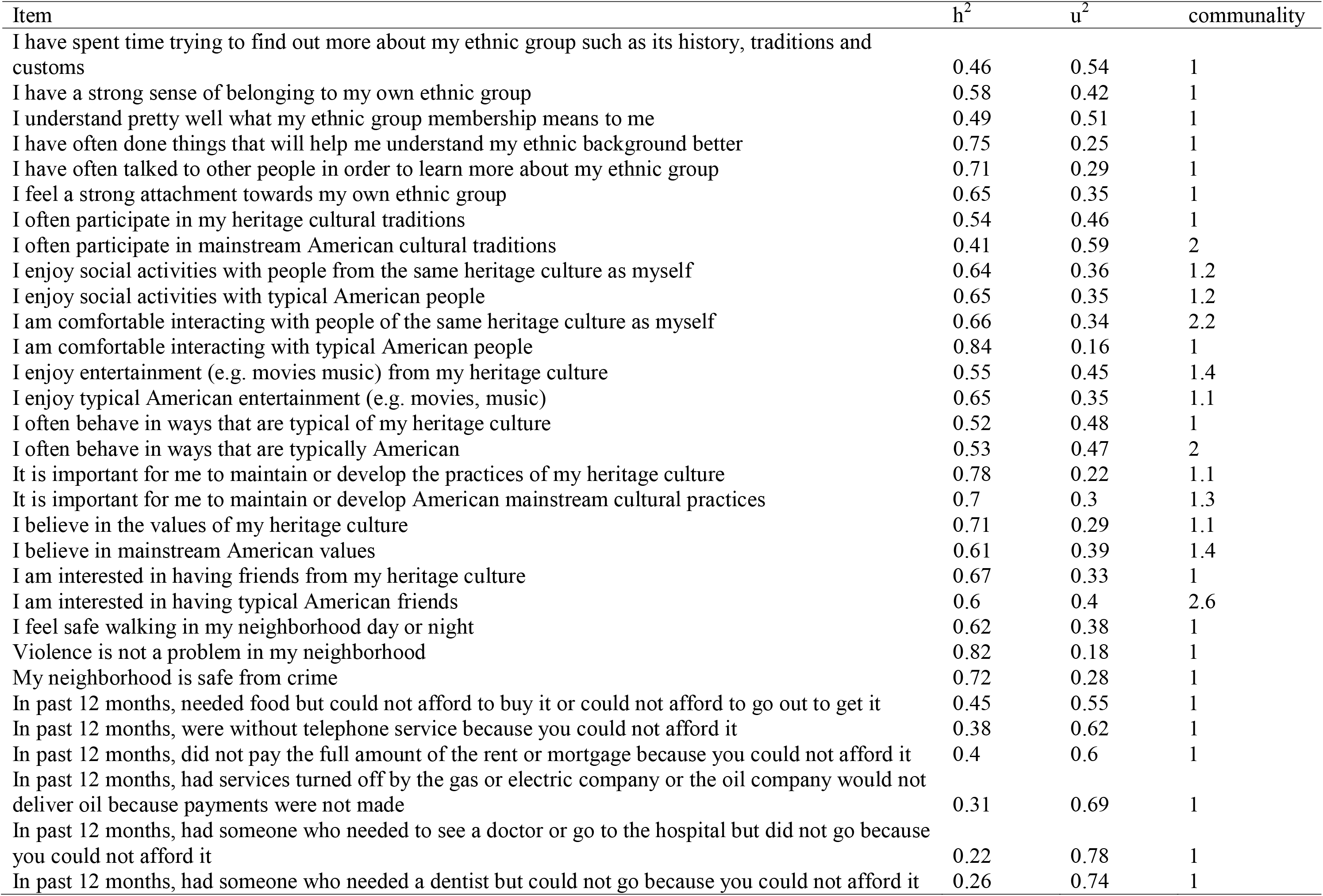
Exploratory factor analysis uniqueness and communality metrics.

Factor loadings were largely consistent with theoretical predictions. Factor 5 was theoretically consistent with the *deprivation* domain. Factor 3, in turn, was theoretically consistent with the *stimulation* domain. The *discrepancy* domain comprised the remaining 4 factors. Factors 1 and 4 index participation in heritage and American culture respectively. Factors 2 and 6, in turn, index wish to understand and/or maintain ethnic group and American cultural practices respectively. Factors were named according to corresponding items and theoretical hypotheses. Factor 1 was named “heritage culture participation”; Factor 2 was deemed “Sense of belonging with ethnic group”; Factor 3 was named “Neighborhood safety”; Factor 4 was deemed “American culture participation”; Factor 5 was deemed “Deprivation”; Factor 6 was named “Maintain American cultural practice.”

### Associations between self-report factors and PLEs

Greater endorsement of the “deprivation” factor/*deprivation* domain related to greater PLEs (*r* = 0.12). Similarly, endorsement of safer neighborhoods/*stimulation* domain related to less PLEs (*r* = -0.09), consistent with predictions. The “heritage cultural participation” and “maintain American cultural practice factors were not found to significantly relate to PLEs. Consistent with predictions, the two factors most relevant to the *discrepancy* domain (“sense of belonging with ethnic group” and “American culture participation”) related to PLES such that greater sense of belonging with ethnic group (*r* = -0.05) and greater American culture participation (r = -0.05) predicted less PLEs (Table 4). The strength of the correlations was subsequently compared to gauge the relative strength of observed associations; the “deprivation” factor association with PLEs was significantly greater than that of the *discrepancy* domain “American culture participation” and “sense of belonging with ethnic group” factors, which survived Bonferroni correction (Table 5).

**Table 4.**
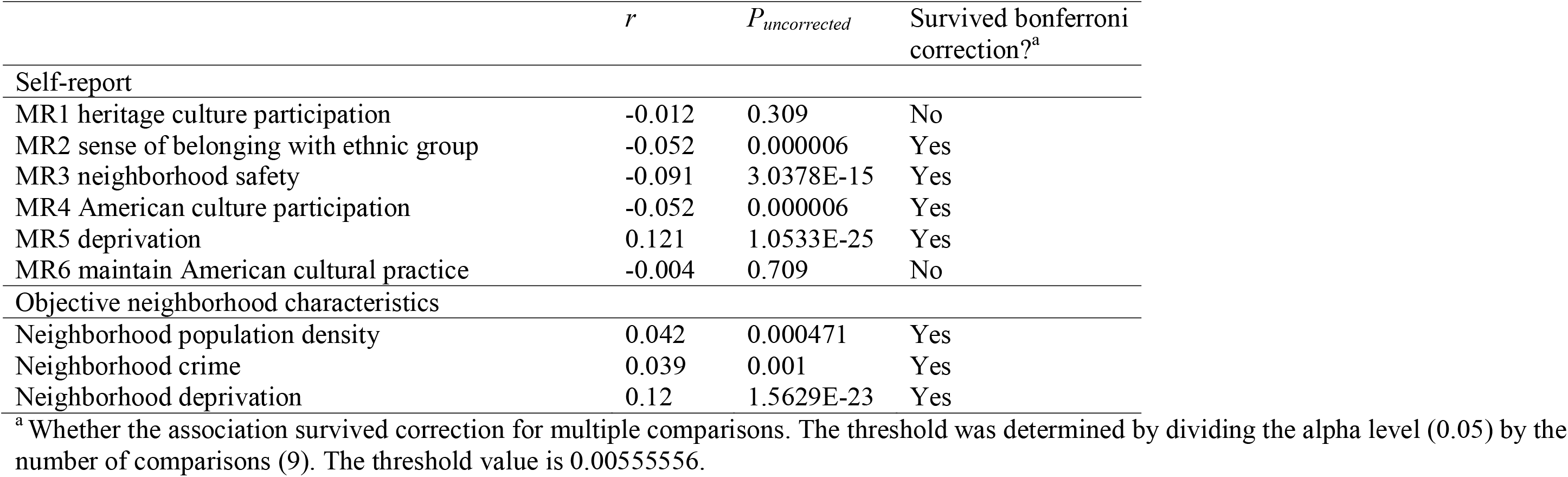
Spearman partial correlations for self-report and objective measures controlling for sex and age. MR3 (neighborhood safety), neighborhood population density and crime are part of the *stimulation* domain. MR2 (sense of belonging with ethnic group) and MR4 (American culture participation) comprise the *discrepancy* domain. MR5 (deprivation) and neighborhood deprivation constitute the *deprivation* domain.

### Associations between objective neighborhood features and PLEs

With regards to objective neighborhood measures, increased *stimulation* domain neighborhood population density (*r* = 0.04) and total crimes (*r* = 0.04), along with *deprivation* domain neighborhood deprivation (*r* = 0.12) all related to increased experience of PLEs (Table 4). The strength of the correlations was compared; *stimulation* domain neighborhood population density and total crimes did not significantly differ in strength of the association with PLEs. However, the association between *deprivation* domain neighborhood deprivation and PLEs was significantly stronger than the associations observed for both *stimulation* domain total crimes and population density (Table 5).

**Table 5.** Comparison of strength of association. MR3 (neighborhood safety), neighborhood population density and crime are part of the *stimulation* domain. MR2 (sense of belonging with ethnic group) and MR4 (American culture participation) comprise the *discrepancy* domain. MR5 (deprivation) and neighborhood deprivation constitute the *deprivation* domain.

| Comparisons | \|Z\| score | $P_{uncorrected}$ | Survived bonferroni correction?<sup>a</sup> |
| --- | --- | --- | --- |
| Self-report comparison |  |  |  |
| MR2/MR3 | 2.446 | 0.01445 | No |
| MR4/MR3 | 2.636 | 0.00839 | No |
| MR5/MR3 | 2.12 | 0.03363 | No |
| MR2/MR4 | 0 | 1 | No |
| MR2/MR5 | 4.376 | 0.00001 | Yes |
| MR4/MR5 | 4.517 | 0.000006 | Yes |
| Self-report/objective measure comparison |  |  |  |
| MR5/ neighborhood deprivation | 0.071 | 0.94318 | No |
| MR3/ crime | 3.55 | 0.00038 | Yes |
| Objective measure comparison |  |  |  |
| Pop density/ crime | 0.236 | 0.81 | No |
| Pop density/ neighborhood deprivation | 4.776 | 0.000002 | Yes |
| Crime/ neighborhood deprivation | 5.344 | <0.000001 | Yes |

<sup>a</sup> For self-report comparisons, the bonferroni-corrected threshold was 0.00833. For self-report/objective measure comparisons, the bonferroni-corrected threshold was 0.025. For objective measure comparisons, the bonferroni-corrected threshold was 0.01667.

### Exploratory comparison of objective and self-report measures

Finally, as an exploratory aim self-report measures and their corresponding objective measures were compared in terms of strength of the association observed. For the *deprivation* domain, the self-reported “deprivation factor”-PLE association did not significantly differ in strength from the neighborhood deprivation-PLE association. However, for the *stimulation* domain, the self-reported “neighborhood safety”-PLE association was significantly stronger than the association observed between neighborhood total crimes and PLEs.

## Discussion

The current study used a large representative sample to examine whether environmental exposures could be distinguished as theorized in the SDD model of psychosis. Further, environmental stressor exposures (self-report and objective) were explored in relation to PLEs. Although we expected three factors to emerge, the EFA of self-report data identified 6 factors, corresponding to heritage culture participation, sense of belonging with ethnic group, American culture participation, neighborhood safety, deprivation, and maintaining American cultural practice. Though more factors were fit than anticipated, these factors did largely fit within the SDD model. Sense of belonging with ethnic group and American culture participation were most consistent with the *discrepancy* domain, which comprises exposures conferring feelings of social exclusion and lack of belonging. Neighborhood safety was theoretically consistent with the *stimulation* domain (along with objective neighborhood total crimes and population density), concerning itself with exposures related to lack of safety and large attentional demands. Lastly, deprivation was consistent with the SDD’s *deprivation* domain (along with objective neighborhood deprivation), constituting exposures related to lack of developmentally appropriate environmental enrichment. Critically, the present investigation is among the first to compare relative strength of the association between these distinct domains of environmental exposure to chronic stress and psychosis risk. Environmental exposure to chronic stress across the three domains related to PLEs: the “deprivation” factor association with PLEs was significantly greater than that of the “American culture participation” and “sense of belonging with ethnic group” factors, suggesting deprivation exposures could relate more strongly to psychosis risk. Consistent with this interpretation, in terms of objective neighborhood measures, the association of neighborhood deprivation and PLEs was also significantly stronger than the association for neighborhood population density or neighborhood crime. Taken together, results lend support to the SDD theory of psychosis, possibly informing etiological models and public policy conceptualizations of prevention and intervention.

EFA was utilized for self-report scales measuring constructs applicable to the SDD theory. As expected, items taken from the NSC survey loaded onto a “Neighborhood safety” factor^53^. These items reflected feelings of safety in one’s living environment, consistent with the *stimulation* SDD domain. Items inquiring about income/resource availability, indexing degrees of environmental enrichment, as consistent with the *deprivation* domain, loaded onto a “deprivation” factor. Lastly, Items related to culture separated into factors relating to “heritage culture participation”, “American culture participation,” “maintain American cultural practice,” and “sense of belonging with ethnic group”. Of these factors, “sense of belonging with ethnic group” and “American/heritage culture participation” are conceptually most consistent with the *discrepancy* domain, as they index current feelings of belonging and participation (as opposed to wish to maintain or participate in American cultural values, as the “maintain American cultural practice” factor captures). Both self-report measures and objective neighborhood features were examined to enrich our conceptualization of the 3 domains and their relation to psychosis risk.

Neighborhood total crimes and population density were part of the *stimulation* domain along with the “Neighborhood safety” self-report factor discussed above. Neighborhood deprivation formed part of the *deprivation* domain, as did the self-report “deprivation” factor.

Self-report and objective environmental exposures were associated with PLEs. Decreased reports of neighborhood safety (along with greater neighborhood total crimes and higher neighborhood population density) predicted greater PLE endorsement. Results are consistent with previous investigations on adolescents and adults finding *stimulation* exposures to relate to increased risk for developing a psychotic disorder ^26,29,31,41^. Further, results suggest that the relation between these environmental exposures and psychosis spectrum symptoms extends to non-clinical psychosis, and is evident as early as late childhood to late adolescence. Mechanisms through which these exposures could be impactful include high attentional demands and feelings of lack of safety—future investigations testing these mechanisms will be crucial.

Increased reporting of deprivation and higher objective neighborhood deprivation predicted greater endorsement of PLEs. Collectively, findings show that both objective and subjective levels of *stimulation* and *deprivation* contribute to the occurrence of PLEs. Reporting greater personal exposure to deprivation showed an association, as did exposure to deprivation in the broader, neighborhood environment. Findings are consistent with a strong body of literature which suggests that exposure to both individual and neighborhood deprivation can confer risk for psychosis^27,41,71-74^, along with adversely affecting physical health and functional outcomes^44,45,75-84^. Future investigations honing in on windows of exposure and differential effects on neurodevelopment will further enrich this line of work.

Of the self-report factors relevant to the *discrepancy* domain, increased “sense of belonging with ethnic group” and “American culture participation” related to less endorsement of PLEs. Observed associations are congruent with evidence that high ethnic density^85-87^ and social cohesion ^88^ can serve as protective factors. Results extend the existing literature by providing evidence that exposures conferring social exclusion and lack of belonging could relate to PLEs as early as late childhood to early adolescence^72,85-91^. “Heritage culture participation” and “maintain American cultural practice” factors, on the other hand, were not associated with PLEs. The lack of significant association between “heritage culture participation” and PLEs could be due to insufficient factor specificity with regards to feelings of belonging. That is, one could participate in one’s heritage culture, and yet not feel a sense of belonging with their surroundings more broadly, or with the majority culture. Psychosis environmental risk factors theorized by the SDD theory *discrepancy* domain include intermediary mechanisms conferring lack of belonging or social exclusion, as well as psychosis risk, such as ethnic minority status^72,86^, low ethnic density^85,87^, and social fragmentation^89-91^. Perhaps the “heritage culture participation” factor does not fully capture these experiences of social exclusion or lack of belonging. “American culture participation,” on the other hand, did relate to PLEs. The association makes sense as American culture in the United States would comprise the “majority” culture. Given the vast evidence of low ethnic density and minority status conferring psychosis risk^72,85-87^, perhaps “American culture participation” indexes comfortability within the majority culture, which could directly impact overall social capital and sense of social cohesion, effecting psychosis risk through this mechanism^32,33,88,92^. The factor “Maintain American cultural practice,” on the other hand, reflects a wish to maintain the cultural practice, not necessarily indexing current participation. Given that it does not index current participation in American culture, this aids interpretation of why the association with PLEs was not observed with this factor. Future investigations assessing other environmental exposures theorized to fall under the *discrepancy* domain and further establishing specificity could aid in offering more nuance to current results.

The literature thus far has been limited in comparing the magnitude of associations between distinct environmental exposures and psychosis risk. The three domains showed differential associations the PLEs, demonstrating their relative contribution could be informative to consider, as well as highlighting the impact of individual domains. In the current sample, observed associations between *deprivation* (objective and self-report) exposures and PLEs were significantly stronger than associations between PLEs and *discrepancy* exposures, with effect sizes for *deprivation* being twice as large. The observed difference is consistent with animal and human literature suggesting that lack of neurodevelopmentally appropriate enrichment can have widespread consequences, impacting a host of key systems necessary for general functioning, as well as overall health and well-being ^44,45,75-84^. Future investigations will be necessary in order to clarify possible mechanisms through which *deprivation* exposures could be particularly relevant for psychosis risk. Further understanding mechanisms for each domain would aid in clarifying and interpreting the extent of their associations with psychosis vulnerability. PLEs are complex, with a multitude of putative contributing factors, and so observed effects, though small, are meaningful when considered in context^93^.

An exploratory aim sought to compare the magnitude of PLE associations between self-report versus objective environmental exposures. The PLE association with self-report deprivation was not significantly different in magnitude than the PLE association with objective neighborhood deprivation. However, this was not the case with *stimulation* exposures. While the PLE association between neighborhood total crimes and population density did not significantly differ in magnitude, the PLE association between neighborhood total crimes and self-report “neighborhood safety” did. Indeed, the PLE association with “neighborhood safety” was more than twice as large as the PLE association with neighborhood total crimes. While preliminary, results suggest that perhaps the association between PLEs and crime exposure is at least partially contingent on conscious awareness: in this case, self-reported lack of safety could represent heightened awareness of the “objective” circumstances, or it could represent inaccurate reporting due to heightened vigilance. Future investigations are needed in order to parse out these possibilities.

Current results are informative for models of psychosis etiology, and emphasize the value of targeting specific environmental factors in preventive health policy efforts for psychotic disorders. Findings aid in refining the SDD theory moving forward. However, these findings must be contextualized within certain limitations. Future investigations may benefit from more fully incorporating other facets of environmental chronic stress exposure, and integrating them within the SDD conceptual framework. Further, future investigations would benefit from exploring neural and biological mechanisms that could underlie the associations observed between environmental exposures and psychosis risk. Collecting information on precise timing of exposure would further add richness to neurodevelopmental conceptualizations of risk and resilience. Lastly, longitudinal investigations would aid our ability to predict directionality of the associations, and account for confounds such as social drift.

## Data Availability

Data is publicly available and can be applied for access on nda.nih.gov

## Acknowledgments

Data used in the preparation of this article were obtained from the Adolescent Brain Cognitive Development (ABCD) Study (https://abcdstudy.org), held in the NIMH Data Archive (NDA). This is a multisite, longitudinal study designed to recruit more than 10,000 children age 9-10 and follow them over 10 years into early adulthood. The ABCD Study is supported by the National Institutes of Health and additional federal partners under award numbers U01DA041022, U01DA041028, U01DA041048, U01DA041089, U01DA041106, U01DA041117, U01DA041120, U01DA041134, U01DA041148, U01DA041156, U01DA041174, U24DA041123, U24DA041147, U01DA041093, and U01DA041025. A full list of supporters is available at https://abcdstudy.org/federal-partners.html. A listing of participating sites and a complete listing of the study investigators can be found at https://abcdstudy.org/Consortium_Members.pdf. ABCD consortium investigators designed and implemented the study and/or provided data but did not necessarily participate in analysis or writing of this report. This manuscript reflects the views of the authors and may not reflect the opinions or views of the NIH or ABCD consortium investigators. The ABCD data repository grows and changes over time. The ABCD data used in this report came from [NIMH Data Archive Digital Object Identifier (DOI) 10.15154/1506121]. DOIs can be found at https://nda.nih.gov/general-querv.html?q=querv=studies%20~and~%20orderBy=id%20~and~%20orderDirection=Ascending.

---

**Figure 1.**
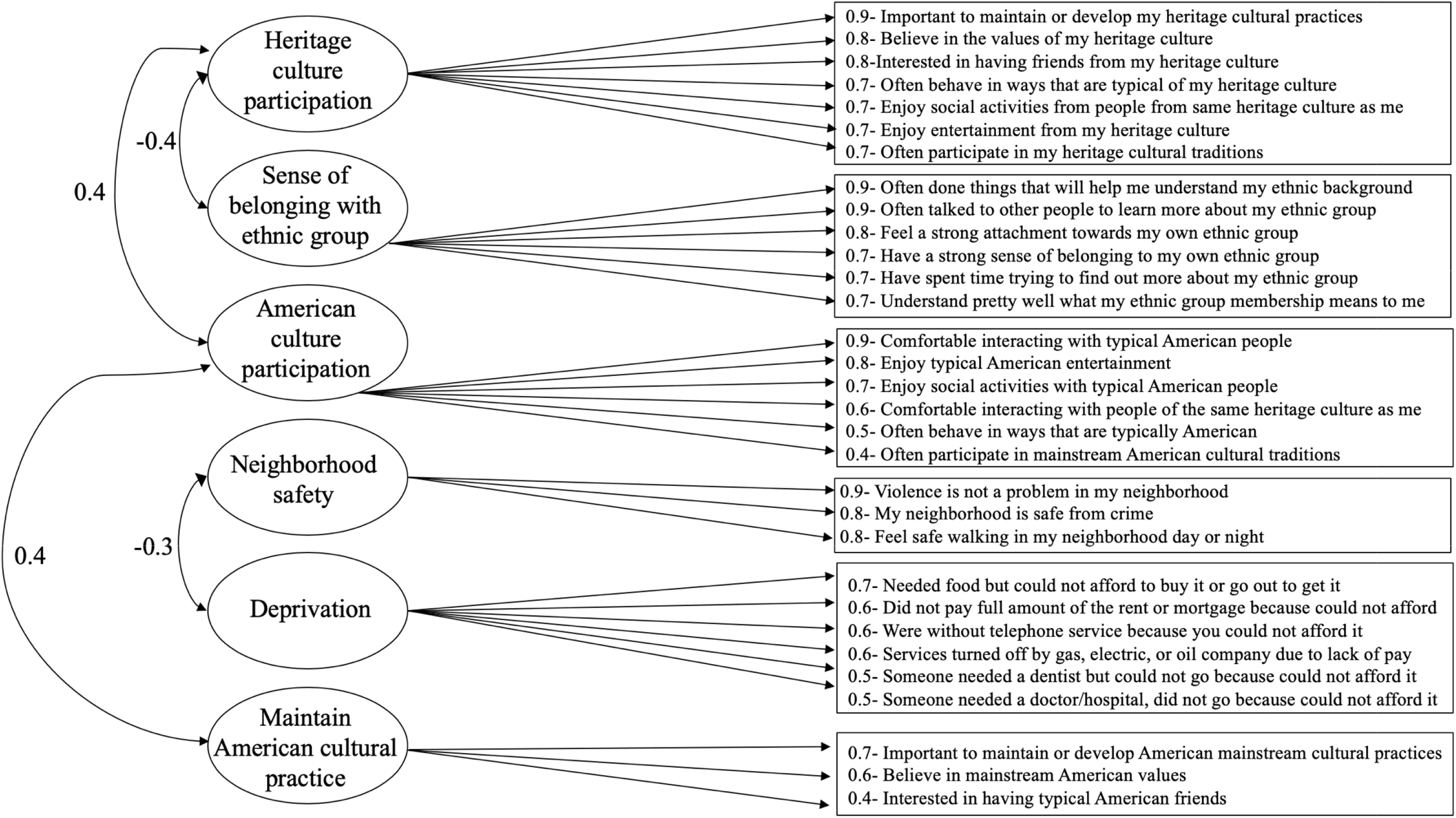
Exploratory factor analysis of self-report items. “Neighborhood safety” is part of the *stimulation* domain. “Sense of belonging with group” and “American culture participation” comprise the *discrepancy* domain. “Deprivation” constitutes the *deprivation* domain. “Heritage c participation” and “maintain American cultural practice” did not relate to PLEs and are not included within the three SDD domains.

**Figure 2.**
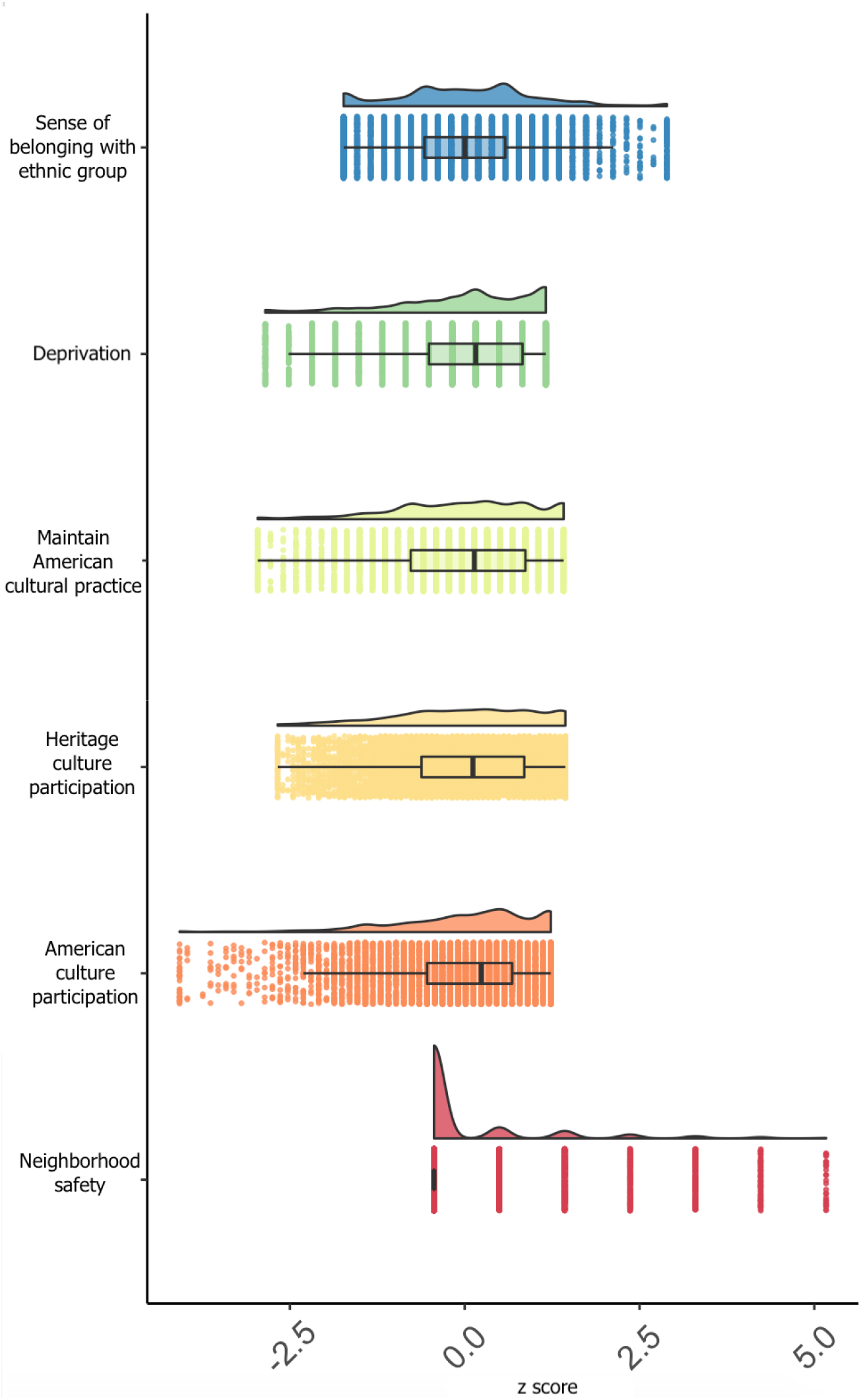
Item endorsement per factor. “Neighborhood safety” is part of the *stimulation* domain. “Sense belonging with ethnic group” and “American culture participation” comprise the *discrepancy* domain. “Deprivation” constitutes the *deprivation* domain. “Heritage culture participation” and “maintain Americ cultural practice” did not relate to PLEs and are not included within the three SDD domains.

**Figure 3.**
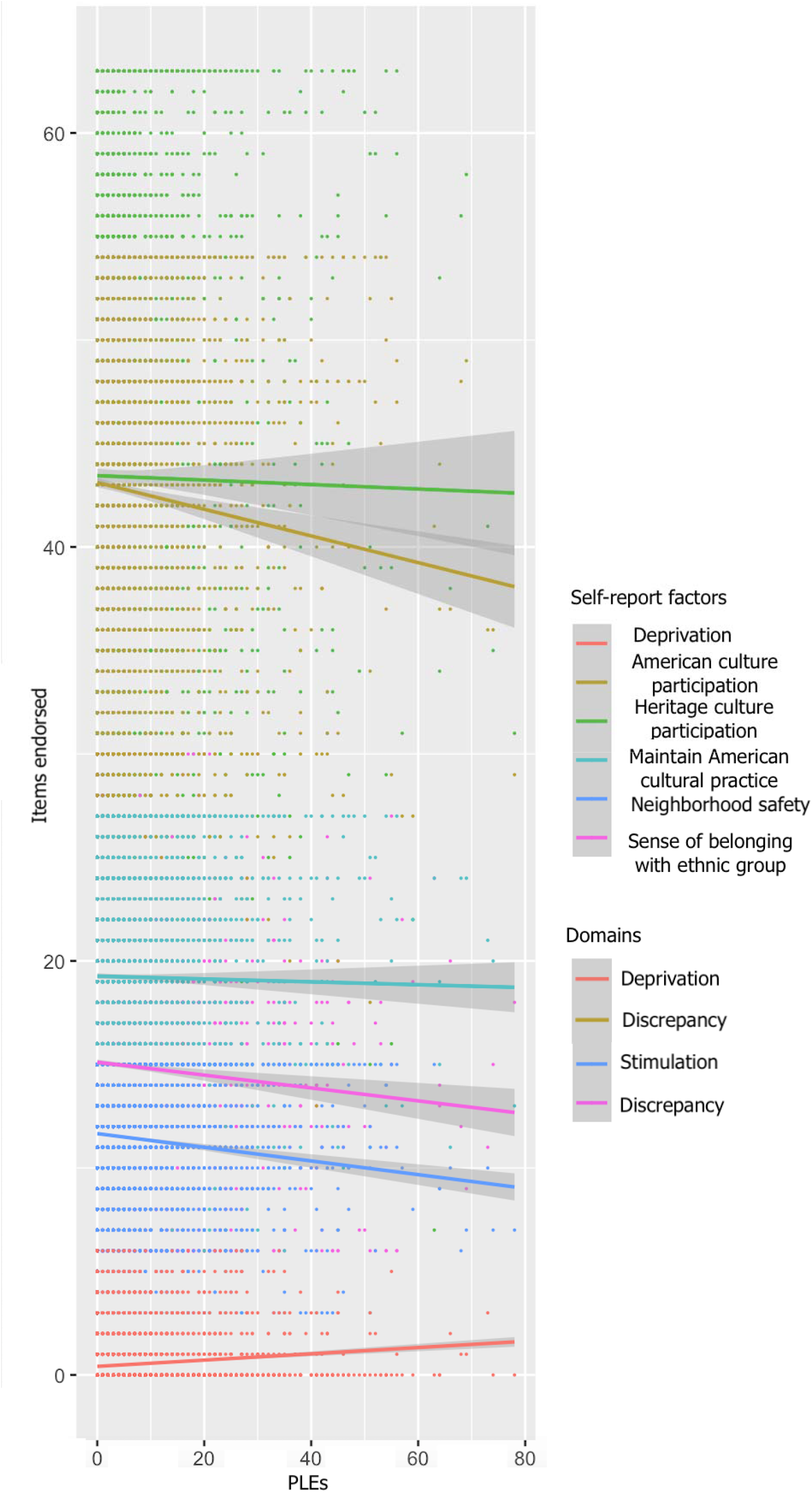
Associations between self-report factors and PLEs.

**Figure 4.**
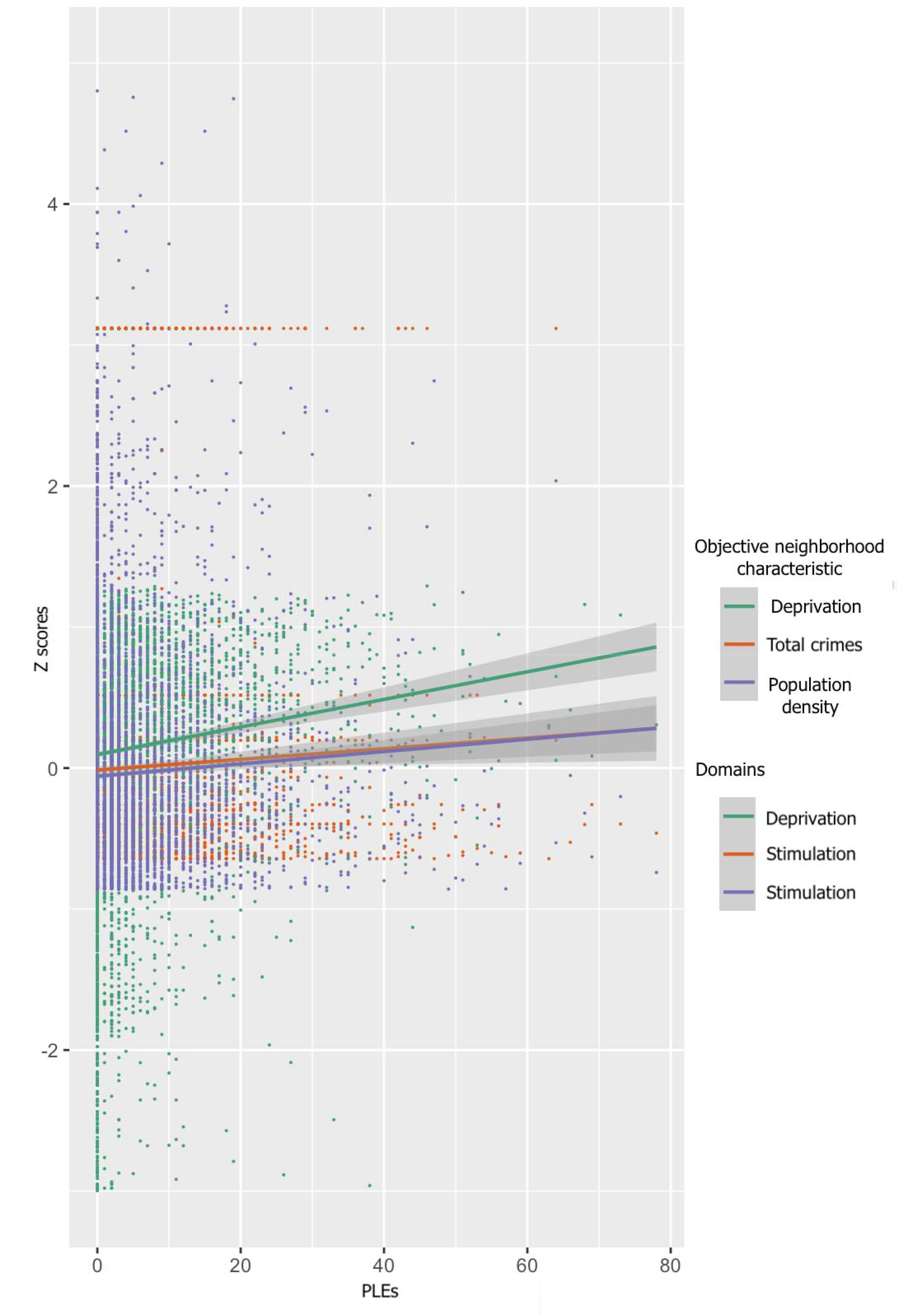
Associations between objective neighborhood characteristics and PLEs.

## Supplementary material

**Supplementary Table 1.**
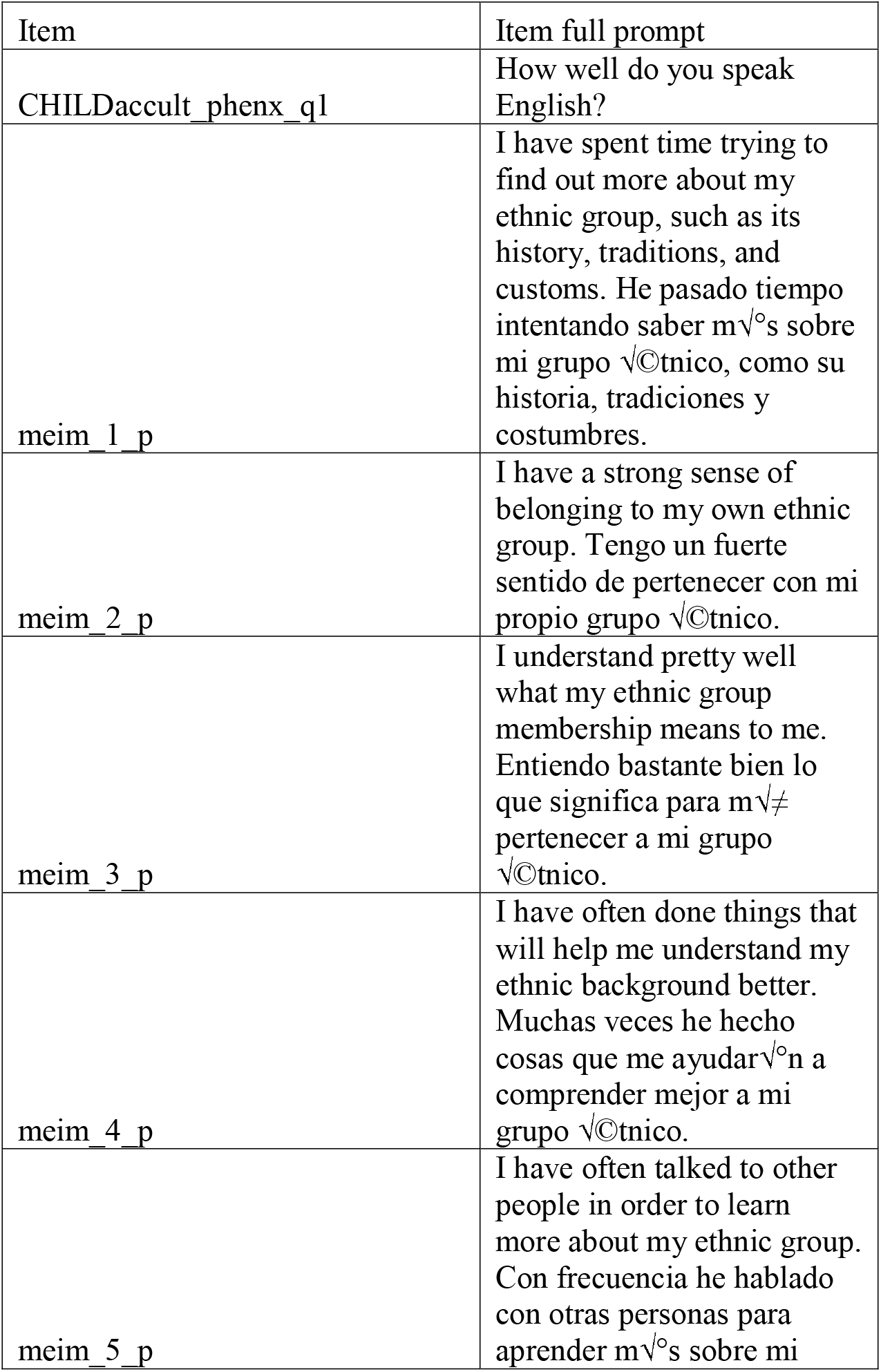
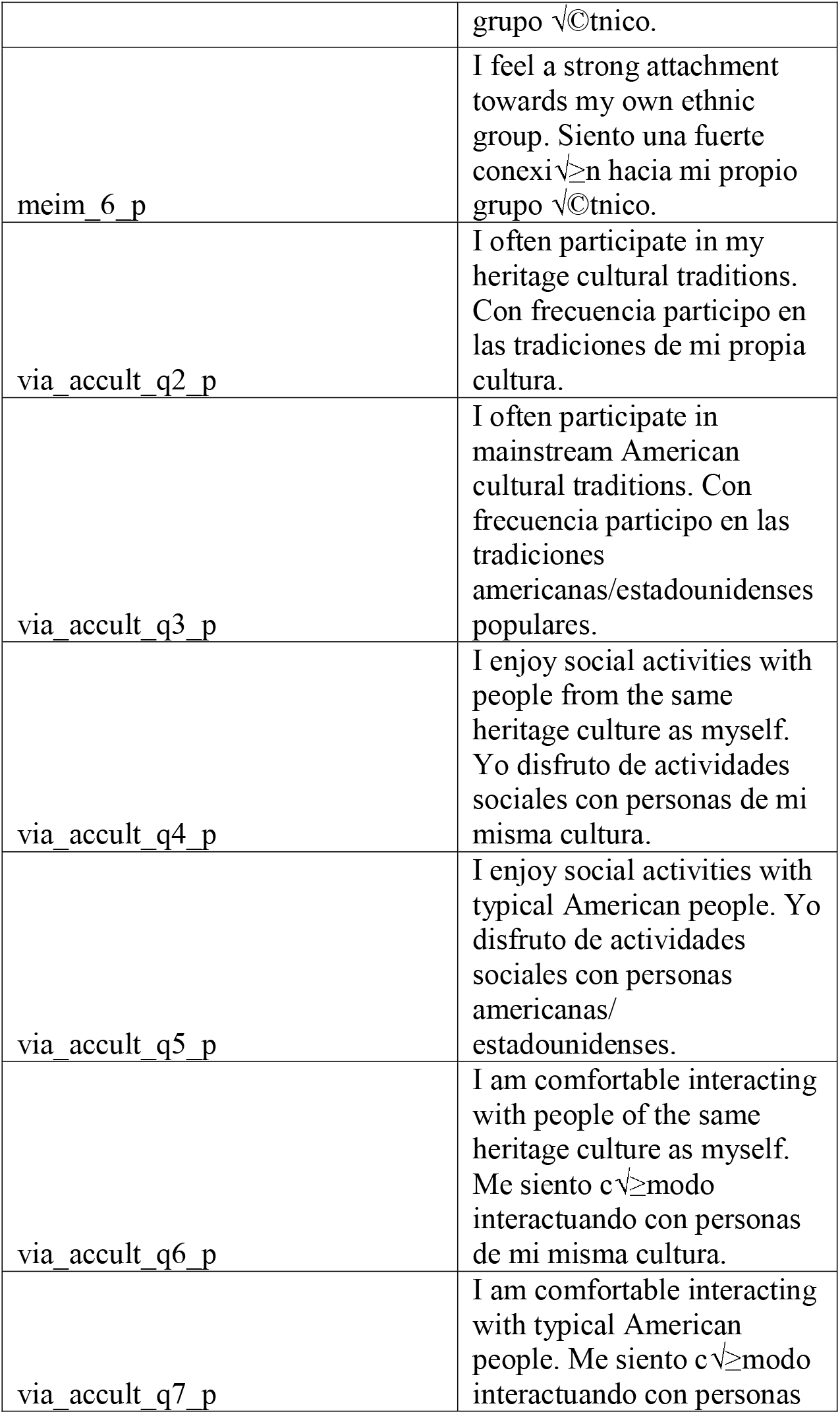
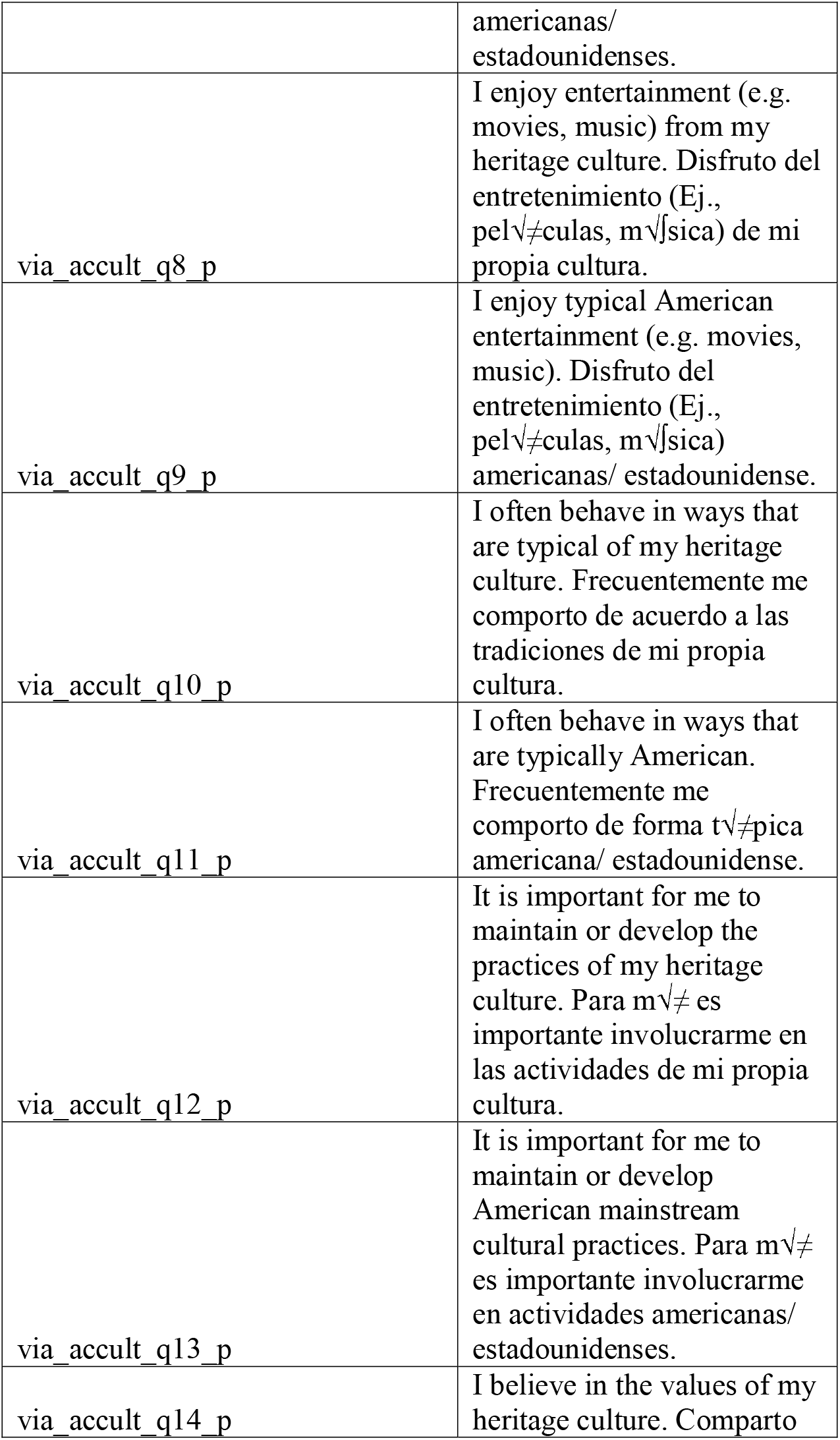
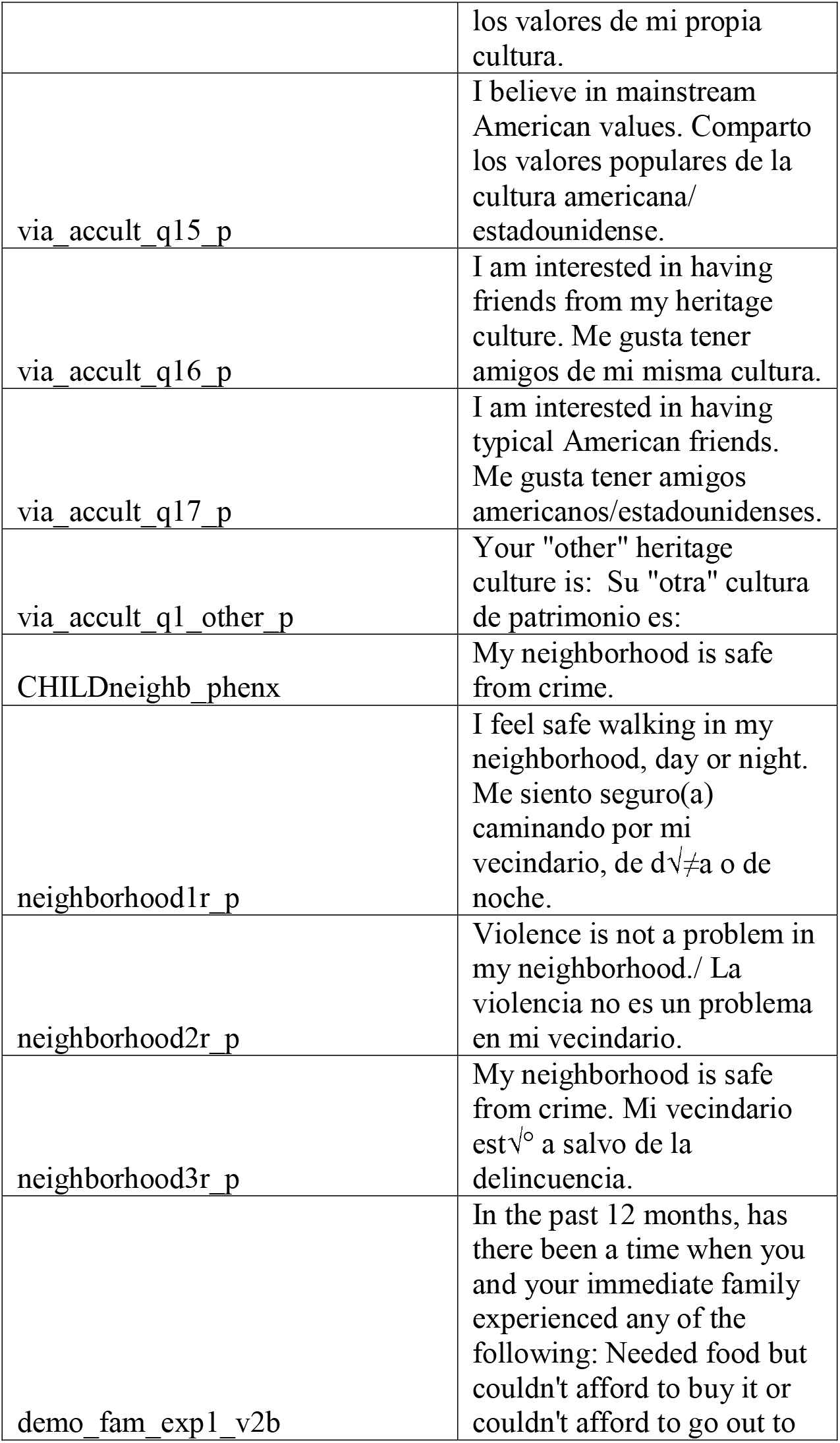
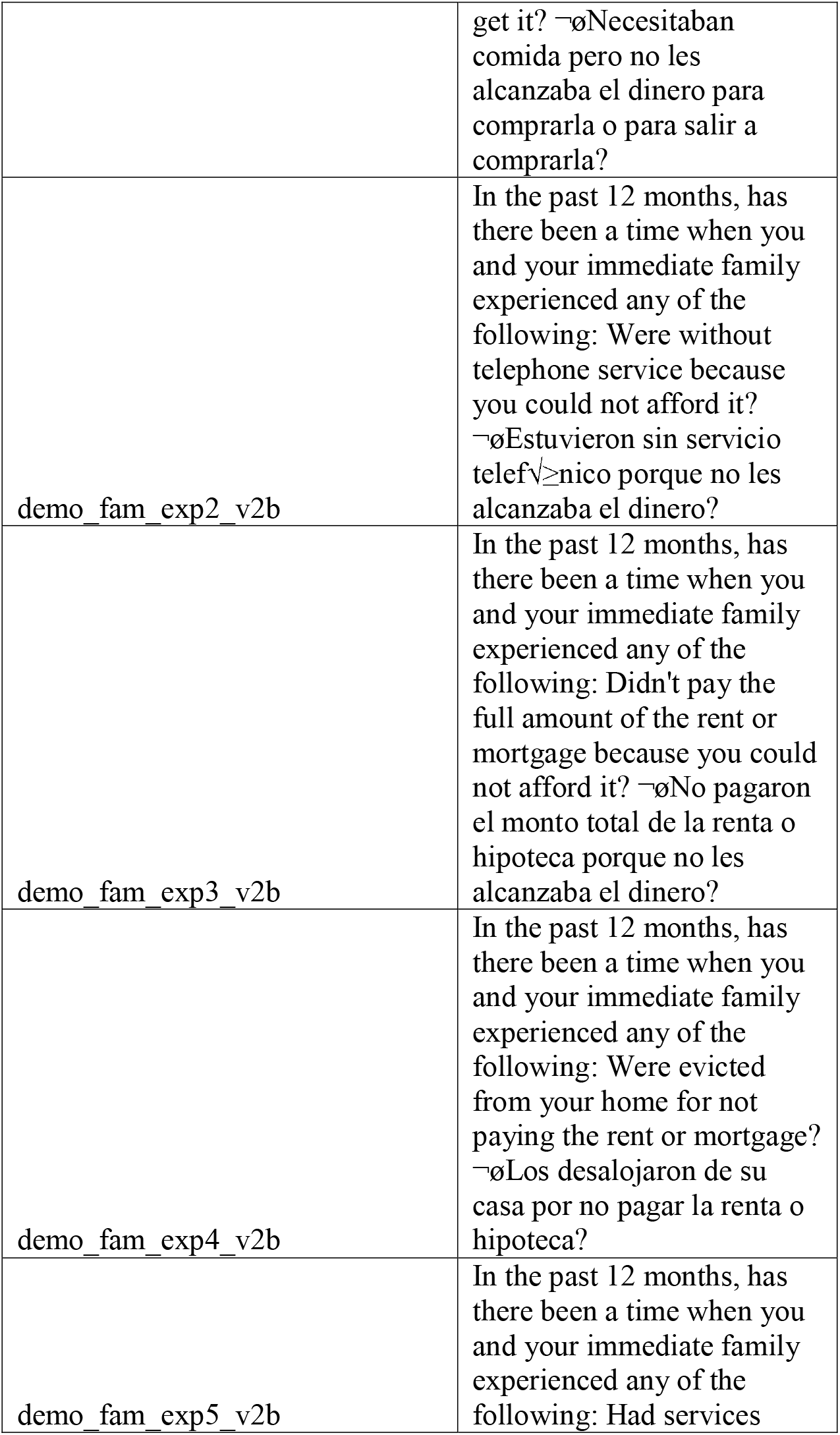
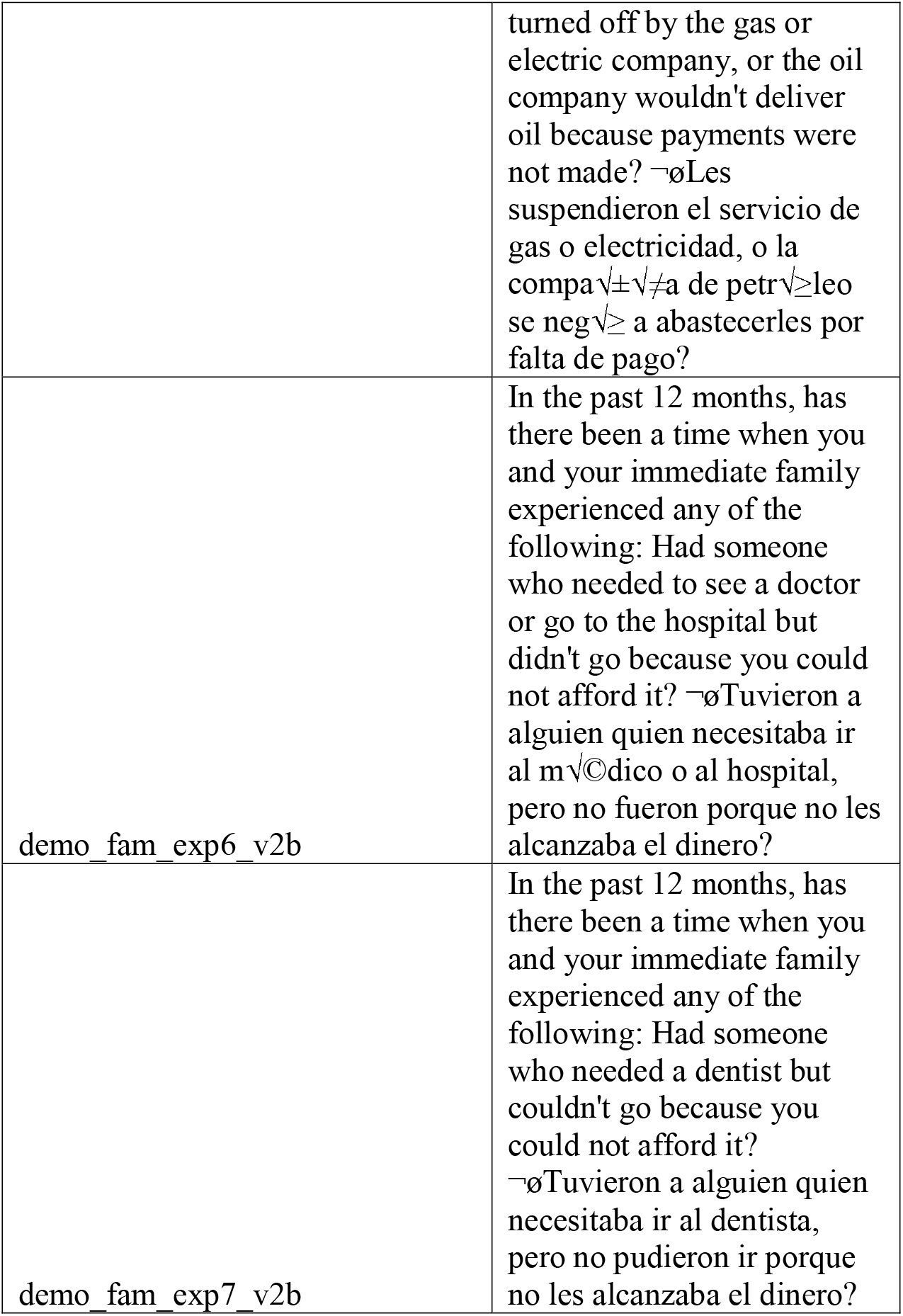
Key including full item prompts.

**Supplementary Table 2.**
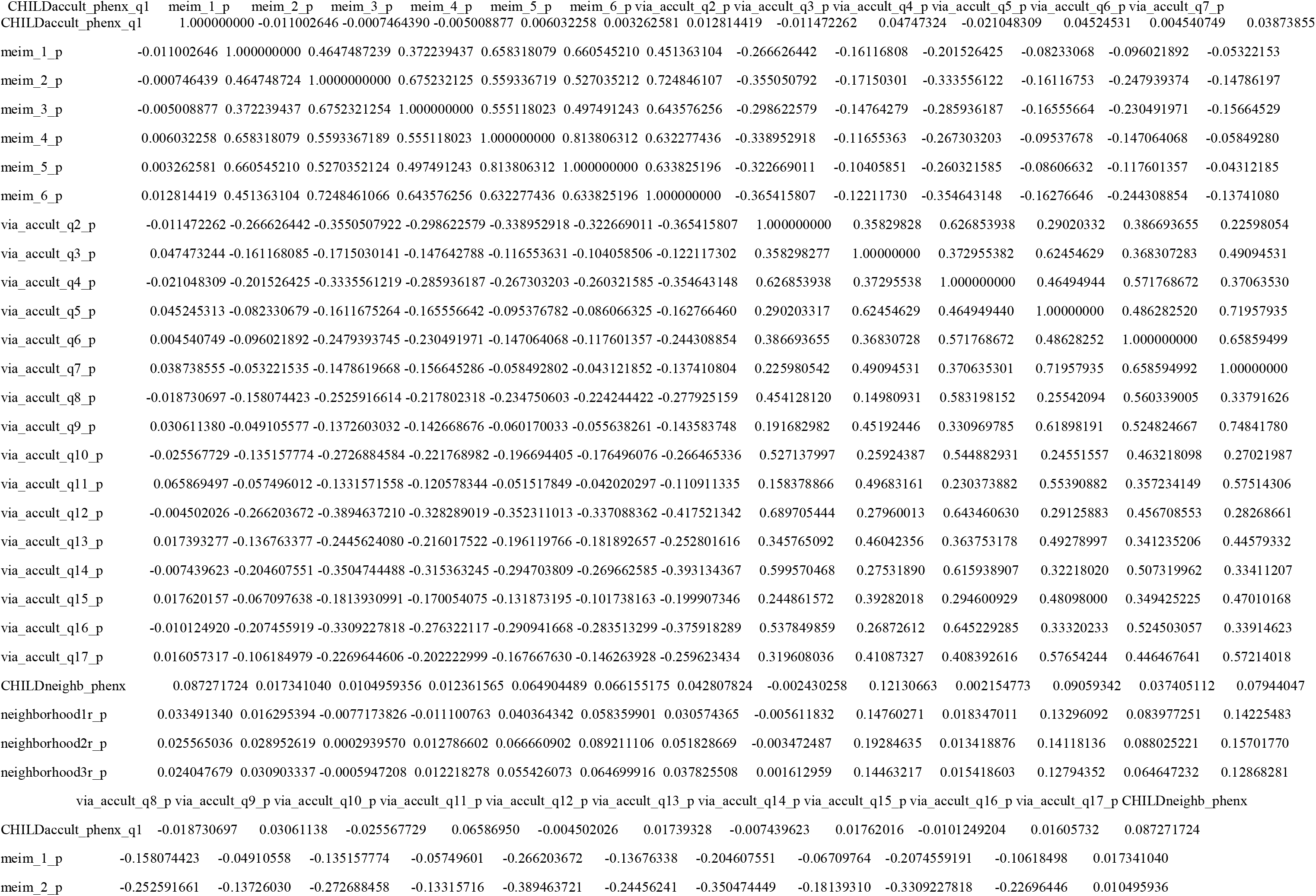
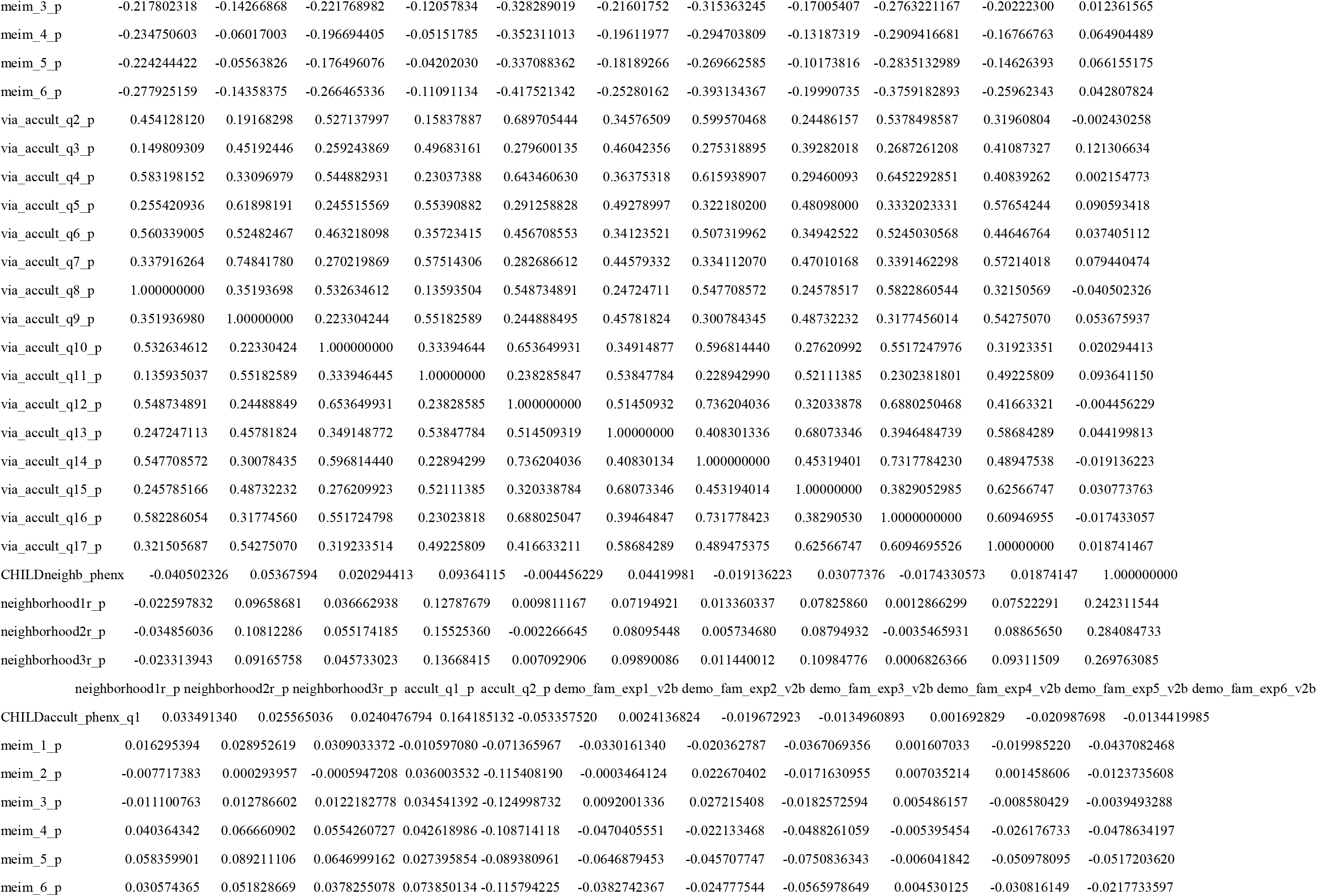
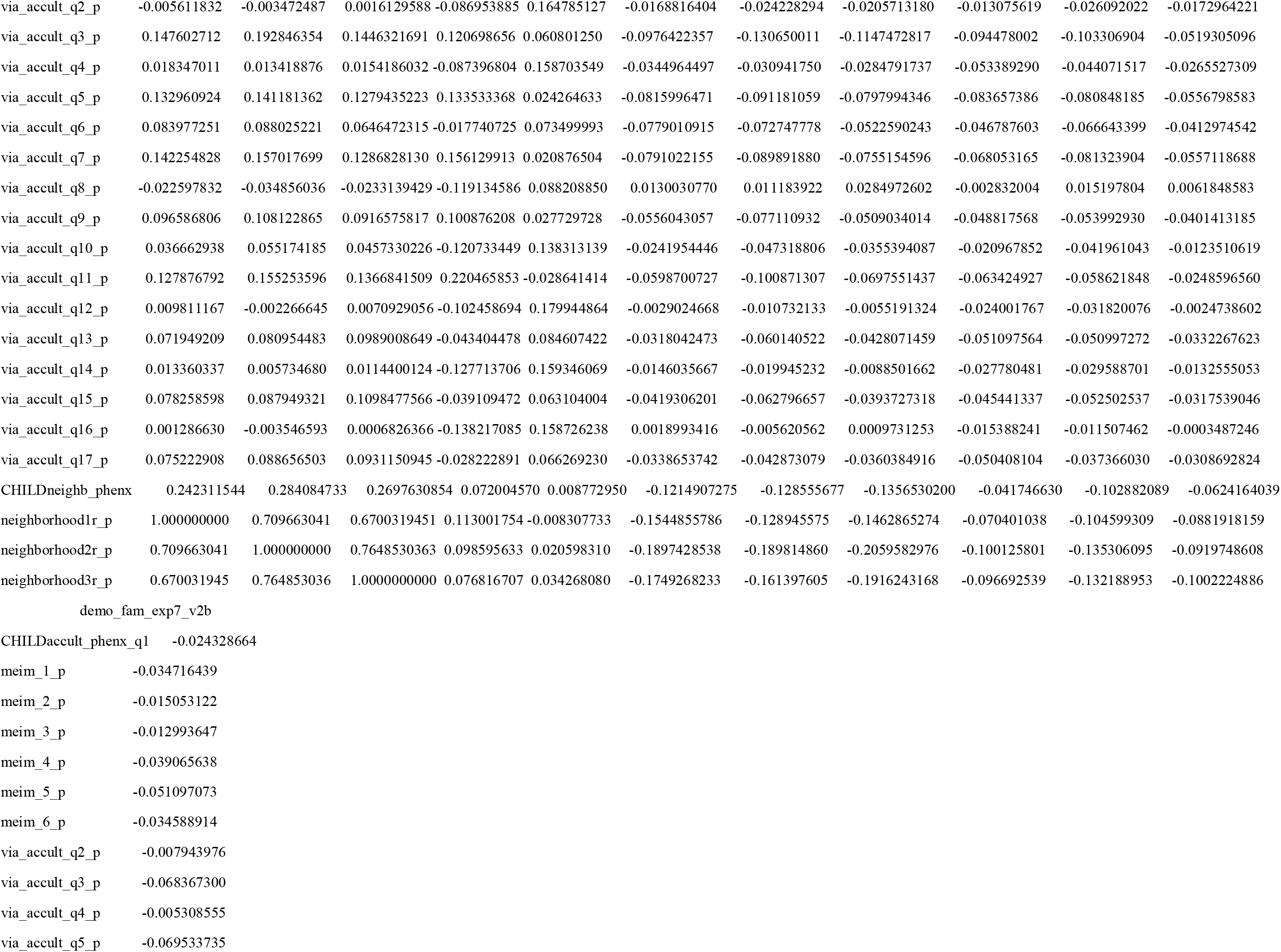
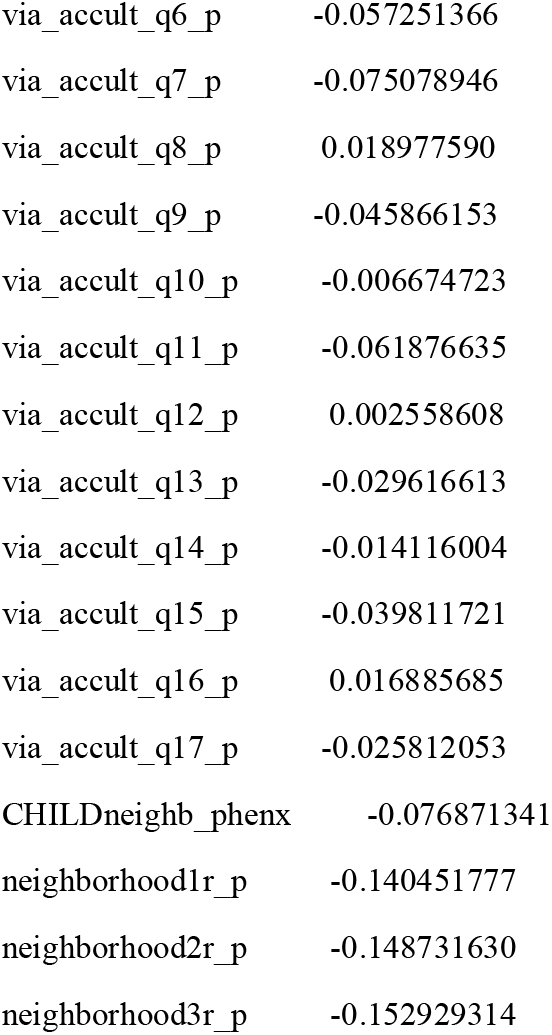
Correlation matrix of self-report items initially considered for exploratory factor analysis.

**Supplementary Table 3.** Exploratory factor analysis factor correlations.

|  | MR1 | MR2 | MR4 | MR3 | MR5 | MR6 |
| --- | --- | --- | --- | --- | --- | --- |
| MR1 | 1 | -0.43 | 0.43 | 0.02 | -0.04 | 0.28 |
| MR2 | -0.43 | 1 | -0.12 | 0.05 | -0.05 | -0.16 |
| MR4 | 0.43 | -0.12 | 1 | 0.16 | -0.14 | 0.39 |
| MR3 | 0.02 | 0.05 | 0.16 | 1 | -0.31 | 0.11 |
| MR5 | -0.04 | -0.05 | -0.14 | -0.31 | 1 | -0.07 |
| MR6 | 0.28 | -0.16 | 0.39 | 0.11 | -0.07 | 1 |

**Supplementary Table 4.** Exploratory factor analysis factor loadings and proportion of variance explained.

|  | MR1 | MR2 | MR4 | MR3 | MR5 | MR6 |
| --- | --- | --- | --- | --- | --- | --- |
| SS loadings | 4.66 | 3.72 | 3.45 | 2.2 | 2.07 | 1.8 |
| Proportion Var | 0.15 | 0.12 | 0.11 | 0.07 | 0.07 | 0.06 |
| Cumulative Var | 0.15 | 0.27 | 0.38 | 0.45 | 0.52 | 0.58 |
| Proportion explained | 0.26 | 0.21 | 0.19 | 0.12 | 0.12 | 0.1 |
| Cumulative proportion | 0.26 | 0.47 | 0.66 | 0.78 | 0.9 | 1 |

